# A survey of livestock to identify the presence of potentially human infective trypanosomes in the *Trypanosoma brucei gambiense* endemic districts of north-west Uganda

**DOI:** 10.1101/2025.11.27.25341151

**Authors:** Enock Matovu, Annah Kitibwa, Darwin Bella Okot, Alex Boobo, Charles Wamboga, Steve J Torr, Sylvain Biéler, Paul R. Bessell, Joseph M. Ndung’u

## Abstract

**Background:** Gambian human African trypanosomiasis (gHAT) is a neglected tropical disease caused by *Trypanosoma brucei gambiense* transmitted by tsetse flies (*Glossina*). Humans are the major host and mass screening and treatment of the human population has contributed to the elimination of gHAT as a public health problem in many countries, including Uganda. There is some evidence that animals may act as hosts for *T. b. gambiense* which may undermine efforts to eliminate transmission. We undertook a study to determine whether cattle and pigs were harbouring *T. b. gambiense* in a region of north-western Uganda where efforts to eliminate transmission of gHAT were ongoing.

**Methodology:** Blood was collected from 2,775 cattle and 531 pigs and examined for the presence of trypanosomes by the haematocrit centrifugation technique (HCT). In addition, polymerase chain reaction (PCR) was used to amplify the trypanosomal internal transcribed spacer (ITS1). Those whose ITS-1 results indicated the *Trypanozoon* group (*T. brucei*) were further tested by sub-species specific PCRs for *T. b. gambiense* and *T. b. rhodesiense*. The results were analysed by species of infecting parasite, geographic location and presence or absence of vector control activities in the area.

**Principal Findings:** Among the cattle samples, 2.2% were positive by HCT (95% CI = 1.7% - 2.8%), while 5% were positive by PCR (95% CI = 4.2% - 5.9%), but this varied depending on location with up to 26.5% positive in one village. Among pigs, 3 samples (0.6%; 95% CI 0.2% - 1.6%) were positive by HCT and none were positive by PCR. No samples were found positive for potentially human infective trypanosomes. There was no significant association between trypanosome species and vector control activities (p > 0.05). Based on these results it was concluded with 95% confidence that human infective trypanosomes are not circulating in livestock at a prevalence greater than 1/1000, and when this is augmented with results of previous surveys potentially human infective parasites are not circulating at a prevalence greater than 1/2848. At the level of the trypanosome species, there is no significant difference in the prevalence of species in vector controlled versus non-vector controlled areas (p > 0.05).

**Conclusions:** These results support the evidence base that livestock are unlikely to be an important reservoir for potentially human infective parasites and therefore not a threat to the elimination of Gambian human African trypanosomiasis in Uganda.

**Author summary:** Two subspecies of the same parasite cause human African trypanosomiasis (HAT) known as sleeping sickness. The east African form has a reservoir in livestock and wildlife with spillover to human hosts, whilst the west African form circulates in humans. However there have been some reports of the west African parasite in animals. Uganda is nearing elimination of the west African form, but to ensure elimination it is essential to verify the absence of the parasite in livestock in the endemic area. In this study a comprehensive livestock survey with testing of blood samples did not find the parasite in livestock. Whilst it is practically impossible to verify the total absence of the parasite, this study combined with studies show that the prevalence of the parasite in cattle or pigs is less than 1/2848 making a very unlikely reservoir.

## Introduction

Uganda is endemic for both the acute and chronic forms of human African trypanosomiasis (HAT) caused by *Trypanosoma brucei rhodesiense* and *T. b. gambiense* respectively. Control efforts by the WHO, several NGOs, as well as the national control program have resulted in a continued decline in incidence of both forms of the disease over the past decades [1]. HAT incidence in Uganda has continued to decline, and since 2020 has not registered any *T. b. gambiense* (gHAT) cases, while just a handful of *T. b. rhodesiense* (rHAT) cases have been recorded (Fig S1). In the *T. b. gambiense* endemic districts in north western Uganda, there were deliberate efforts to improve access to HAT screening and case detection [2]. This was alongside vector control activities that have been shown to be effective at reducing vector numbers and gHAT case numbers [3–6]. Thanks to this and other efforts, only 8 cases were reported between 2017 and 2020, and not one case of gHAT has been reported since then (Fig S1).

As a consequence of these efforts, in May 2022 the WHO validated the elimination of gHAT as a public health problem in Uganda [7]. A component of the validation dossier was understanding the role of domestic animals in transmission and in particular their role as potential reservoirs of the disease.

In the *rhodesiense* areas of south-eastern Uganda, it has been well demonstrated that cattle, dogs and pigs are reservoirs of the acute disease [8–10]. In north western Uganda, there had previously been two surveys to investigate animal African trypanosomiasis (AAT) and the role of animals in chronic HAT transmission [11,12]. These surveys did not cover the entire area at risk and neither study was able to demonstrate any potentially infective trypanosomes in cattle, goats, sheep, dogs and pigs [12]. Bloodmeals among the *G. fuscipes* vectors primarily comprise cattle (39%) and human (37%), with smaller numbers of forest cobra and monitor lizard [13]; there were some pig samples which had also been identified in previous studies [8].

The number of livestock in that region is also not as high compared to those in *rhodesiense* endemic areas where some districts are more populated by pastoral communities. The national livestock census of 2008 put the figures in the study area at 643,658 cattle; 1,187,039 goats; 325,694 sheep and 82,668 pigs [14]. Assuming a 3% increase per annum, at the time of study the populations would be 0.89m cattle, 1.60m goats, 0.44m sheep and 111,000 pigs. Subsequent to the fieldwork a livestock census was carried out in 2021 which rather confirms the projected livestock numbers putting cattle at 835,203 cattle, 1,588,277 goats, 373,774 sheep and 105,979 pigs [15]. Secondly, the frequent periods of political instability in South Sudan leads to refugees who cross with animals.

Given this gap in knowledge about the role of livestock as reservoirs of HAT in north western Uganda and the need for information to support elimination of HAT in the region, we undertook a study to determine whether livestock in the region harbour potentially human infective trypanosomes, which might threaten efforts to eliminate transmission of gHAT.

The key objectives of the study were:

1. To identify to species level, the trypanosomes circulating in domestic animals in T. b. gambiense endemic districts of north western Uganda
2. To determine whether domestic animals in that region harbour potentially human infective T. brucei.
3. To identify any association between tsetse control activities and circulating trypanosome species.

## Methods

### Study area

The study area was the gHAT focus in north-western Uganda, which as of 2022 comprised 10 districts (Adjumani, Amuru, Arua, Koboko, Madi-Okollo, Maracha, Moyo, Obongi, Terego and Yumbe) with a population of 2.22 million, and a large and fluid population of refugees from neighbouring Republic of South Sudan [2,16,17]. The area has a large livestock population, dominated by ruminant farming and sporadic pig rearing.

Between August and October 2019, we carried out field surveys targeting selected parishes across the study area covering the 7 districts of Adjumani, Amuru, Arua, Koboko, Maracha, Moyo and Yumbe (district boundaries have subsequently changed but here we use the boundaries as they were at the time of the study in 2019).

### Sample size estimation and site selection

Focusing on cattle and pigs, we estimated a required sample size of 200 animals from each of 16 sampling sites (total of 3,200 animals). This sample size gives 80% confidence (power) of detecting at least one infected animal at a gHAT prevalence of 1/1,000. To estimate the required sample size we:

1. Test a range of number of sampling sites from 1 to 20 (nSites)
2. Randomly sample the prevalence of gHAT at each site from a beta (1, 1000) distribution (cPrev) equivalent to a median prevalence of 1/1000.
3. Test at each sample site a number of livestock samples (nAnimals) from 10 to 500 at increments of 10, based on the cPrev for that site and nAnimals we use a binomial distribution to sample the number of positive cattle at that site
4. Run each combination for 100,000 iterations.
5. Calculate for each combination of nSites and nAnimals the proportion of iterations with one or more positive cattle
6. Find the smallest combination of nSite and nAnimals which gives one or more positive cattle with a probability greater than 80%.

For practical reasons, sampling was done at the parish level. Parishes were selected based on sampling parishes with greater cattle numbers, greater tsetse numbers (from reported tsetse survey data), greater levels of HAT RDT positivity in humans and the number of confirmed HAT cases.

The following sites (parishes) were selected, including six where no tsetse control had been undertaken in the recent past (Fig S2):

Adjumani district: Okangai

Amuru district: Bibia, Pogo

Arua district: Aliba-Robu, Ayuri

Koboko district: Godia, Lurujo, Asunga

Maracha district: Mundru, Robu

Moyo district: Dufile, Gweere, Ewafa,

Yumbe district: Kerwa, Gojuru, Oluba

### Sample collection

Farmers owning cattle and pigs were mobilized through the District Veterinary Officers in consultation with local leaders, to bring their animals at a central site selected per parish. Owners presented with cattle and pigs and where the owners consented animals were bled by venipuncture into EDTA tubes, and the blood subjected to the haematocrit centrifugation technique (HCT) [18] to estimate the parasitological prevalence per site. The blood was centrifuged at 4000rpm for 10 minutes to separate out plasma. Both the plasma and blood cell pellets were frozen in liquid nitrogen; the former for future studies and the latter for DNA extraction and downstream analyses to identify the infecting trypanosome species [19,20].

In the laboratory, the frozen blood pellet was thawed and gently homogenised using a sterile Pasteur pipette. Thereafter, 100µl of the suspension was used to extract DNA using a commercially procured Quick gDNA mini prep kit (Zymo Research, Irvine, CA, USA) following the manufacturer’s instructions. Purified DNA was eluted in 50 µL PCR water and stored frozen at −20 SC until use in the PCR reactions. We started with the PCR targeting the Internal Transcribed Spacer (ITS-1) as previously described, first by the single-step ITS-1 PCR [20] and nested ITS-1 PCR [19] for samples in which no signals could be picked by the single step PCR. The analytical sensitivity of Single step ITS1 PCR was reportedly between 10 pg and 100 pg (100-1000 trypanosomes/ml [20]) while that of a nested ITS1 was 49 pg/ ml which is equivalent to less than a single trypanosome in the analysed sample [19]. Any samples that indicated presence of *T. brucei* were further subjected to the *T. b. gambiense* and *T. b. rhodesiense* specific PCRs targeting the *Trypanosoma gambiense* Surface Glycoprotein (TgsGP) and the Serum Resistance Associated (SRA) genes respectively, using nested PCR as previously described [21], the latter to rule out the spread of the acute rHAT with animal movement from the south-east. The Primers and PCR conditions deployed in this study are indicated in Table 1 below:

**Table 1:**
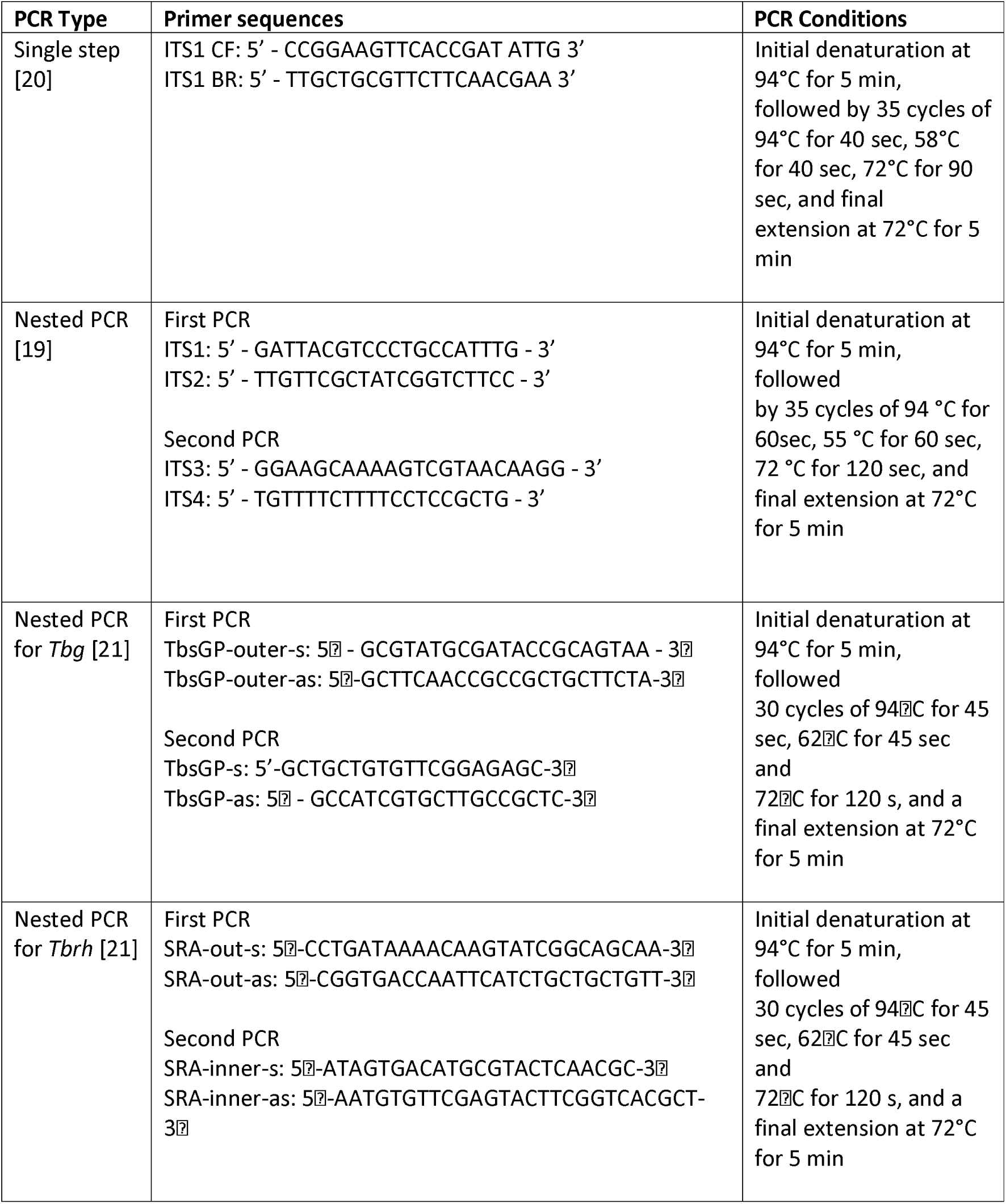
Primers and PCR conditions deployed in this study.

As an incentive to farmers, their trypanosome positive cattle were treated with diminazene aceturate. This drug is also used for treatment of the tick-borne disease babesiosis, thus treatment of the animals was likely to lead to an improved general herd health.

## Data analysis

In the event of the pathogen not being found in these surveys, we estimate our confidence that the parasite is indeed absent and if it were still circulating what would be a realistic prevalence. We evaluated the likelihood of absence of the pathogen by applying a negative binomial distribution to the actual sample size. We simulated a range of prevalences from 1/(100…5000) and used the rnbinom() function in the R statistical environment [22] to give the probability of finding at least one positive sample at a given prevalence and the size of the livestock population that was sampled. To add further weight to the analysis, we augmented our results with data from three previous studies [11,12,23] (Table 2). This brought in an additional 4,320 cattle and 902 pig samples.

**Table 2.** Additional livestock testing data used to augment the survey results from surveys that also found absence of human infective trypanosomes in the area.

| Paper | Sampled years | Districts | Cattle sampled | Pigs sampled | Other animals |
| --- | --- | --- | --- | --- | --- |
| [11] | 2004-2008 | Koboko, Moyo, Arua | 2232 | 161 | 874 |
| [12] | 2011-2014 | Koboko, Arua | 2088 | 400 | 0 |
| [23] | 2019 | Arua and Maracha | 0 | 1262 (341 by PCR which are used for analysis) | 0 |

We also evaluated the difference between test results in areas that had undergone vector control using Tiny Target deployments versus non-vector controlled areas [5]. To do this we used a mixed effects logistic regression model with the test result the outcome and the clustering at village level as a random effect. This was implemented in the lme4 [24] package for R [22]. 95% confidence intervals for binary data were calculated using the Hmisc package for R [22].

## Ethics statement

The study protocol was reviewed and approved by the Ministry of Health (Vector control Division Research Ethics Committee; VCDREC) and final clearance was provided by the Uganda National Council for Science and Technology as study number A604.

## Results

Samples were taken from 2,775 cattle from 16 villages and 531 pigs from 10 villages (Table 3 and Fig S3). By HCT, 60 (2.2%) samples were positive among cattle (95% CI = 1.7% - 2.8%) and three samples from pigs were positive (0.6%; 95% CI 0.2% - 1.6%). By PCR, 140 samples (5.0%, 95% CI = 4.2% - 5.9%) were positive from cattle, but this varied hugely from 26.5% (n=155) in one village (Mindrabe) to 0% in the villages of Esia (n = 116) (Table 3). No pigs were PCR positive (Table 3).

**Table 3.**
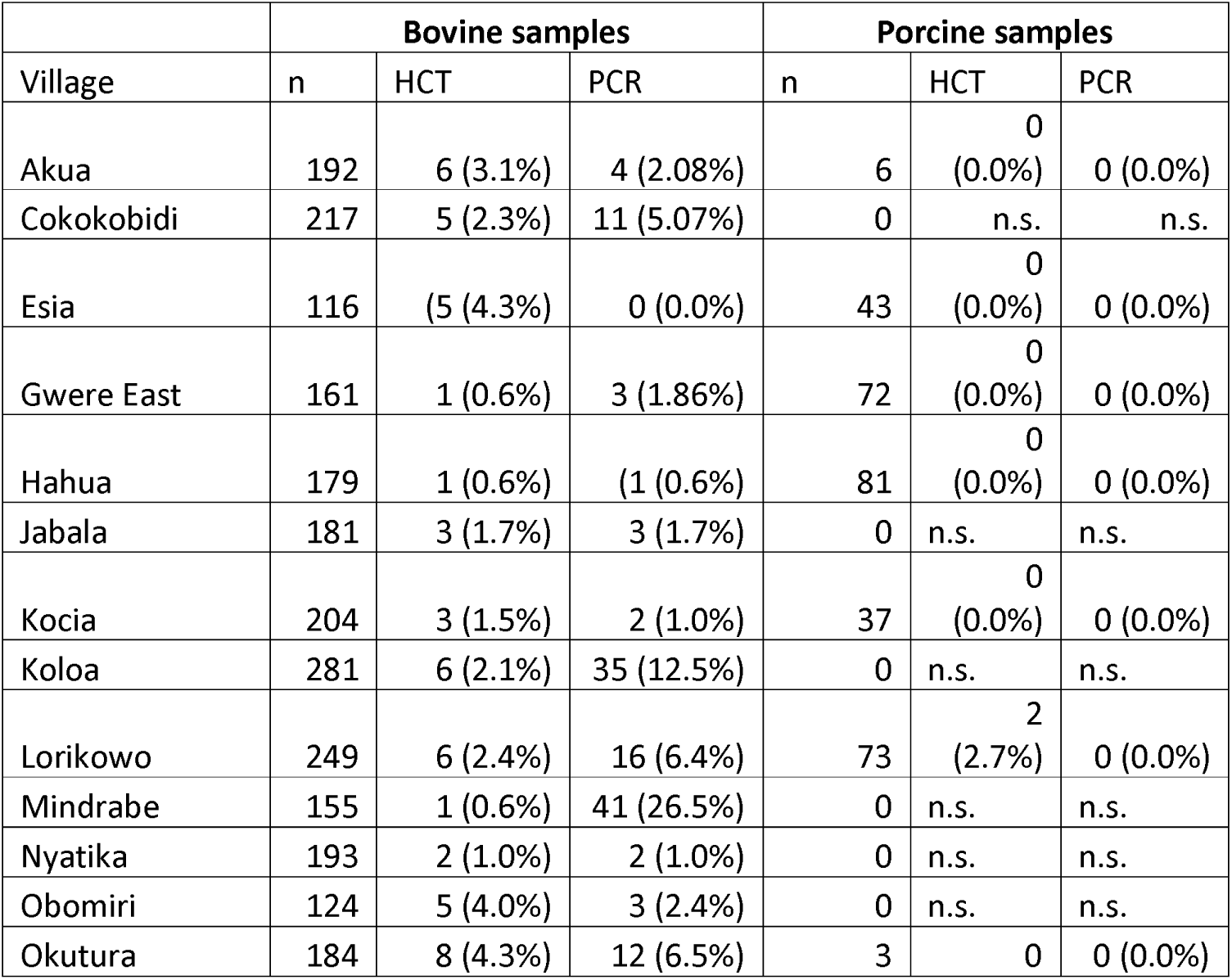
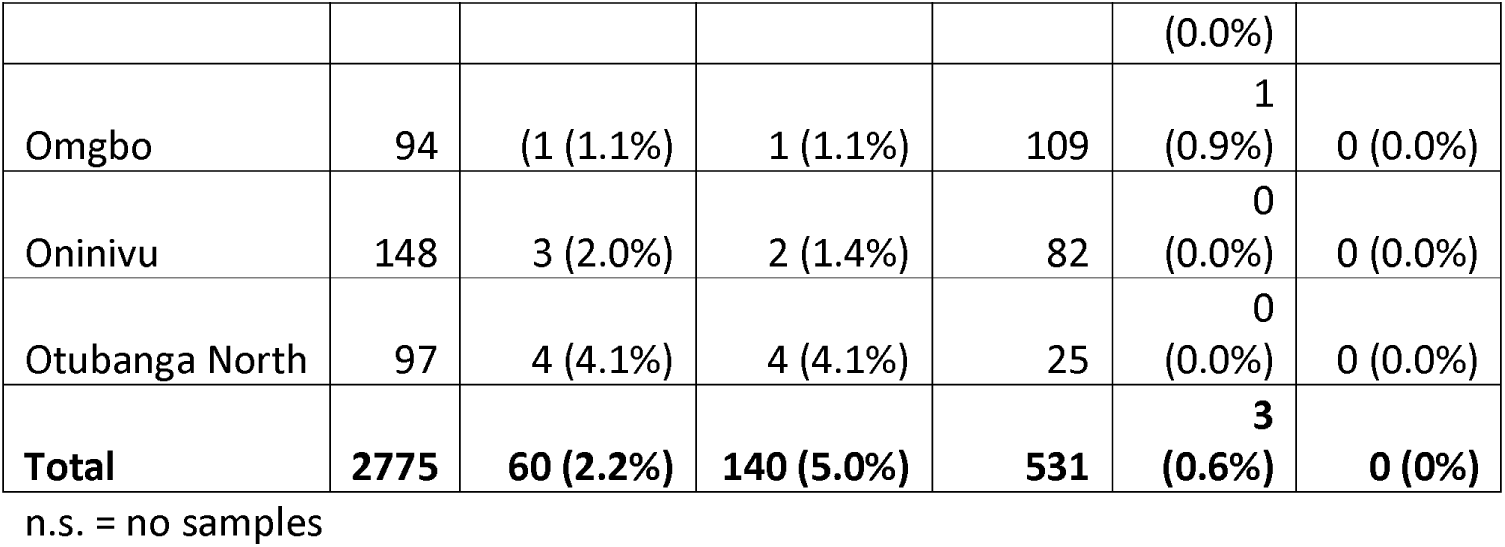
Summary of the key results by parasitology and PCR.

Considering the whole sample collection (3,306 samples from 2,775 cattle and 531 pigs), PCR yielded many more positive results (from 140 samples; 4.2%) than the HCT (63 samples; 1.9%). The species composition among the PCR positive samples was mainly *T. congolense* (47.1%), followed by *T. vivax* (38.6%) and the benign *T. theileri* (29.3%), while *T. brucei* was least represented (12.9%) (Supplementary table S1). Noteworthy is that 27.9% of the positive samples haboured more than 1 infecting trypanosome species in the same animal (Supplementary table S2).

No samples were positive by *T.b. gambiense or T.b. rhodesiense* specific PCR. Whilst it is impossible to measure zero in such an instance, we can conclude with 95% certainty that if human infective trypanosomes had been present among cattle then it would have been at a prevalence below 0.11% and below 0.56% among pigs, and between both species the overall prevalence would be below 0.09%. Based on the number of animals tested, if the pathogen had been circulating at a prevalence of 1/1,000, the probability of picking an infected animal would have been 96.3%. Based on this, we can say with 95% confidence that if human infective trypanosomes were circulating, then the prevalence among all livestock was less than 1/1,104; among cattle, the corresponding prevalence was less than 1/927 and among pigs less than 1/178 (Fig 1).

**Fig 1.**
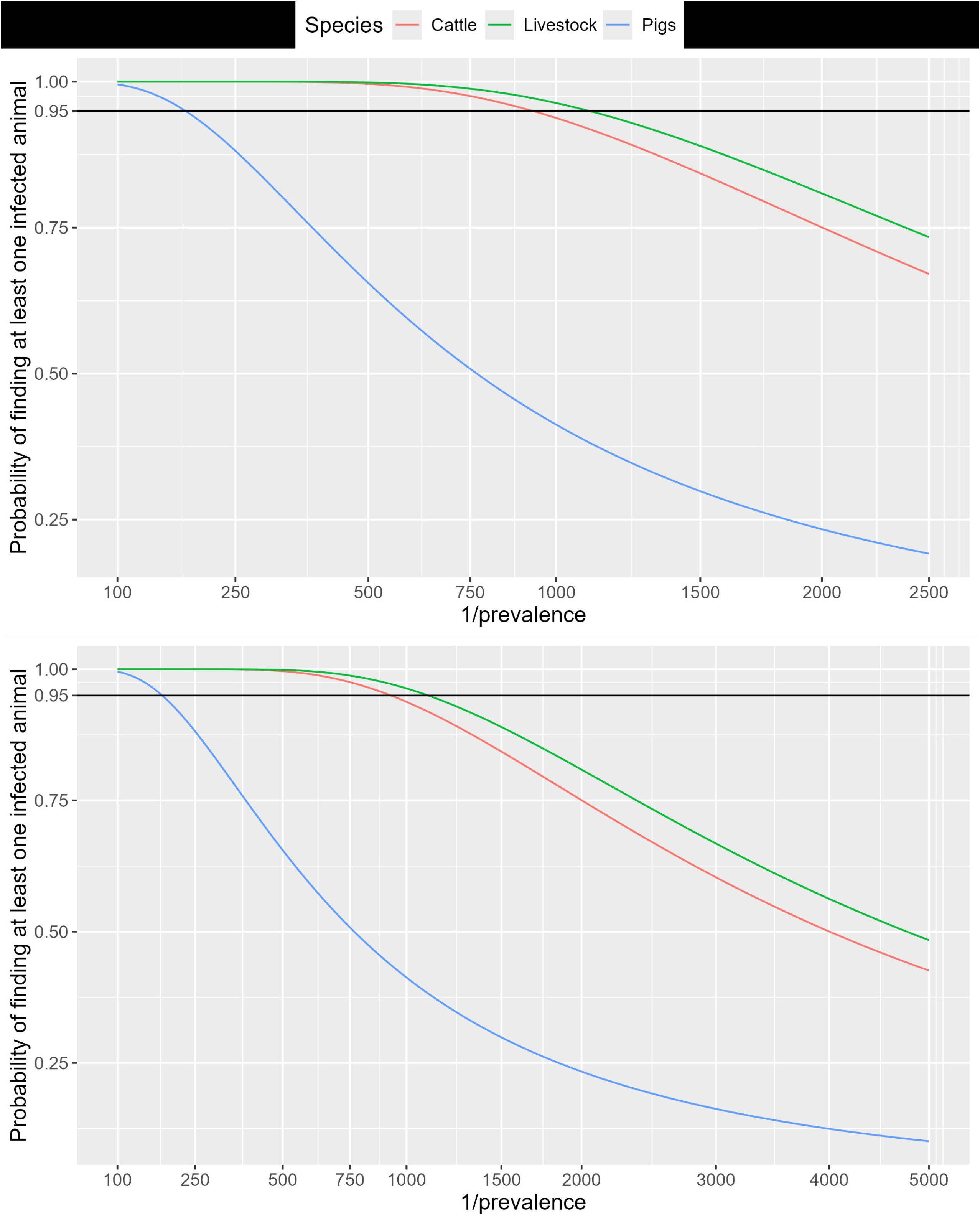
Probability of finding trypanosomes. (Top) Cumulative distribution function of the changes in the probability of finding at least one trypanosome infected animal as the prevalence varies given the sample sizes for cattle, pigs and all livestock. (Bottom) Updating the results from the survey to include results of sampling from previous studies in the area. The lines show how the probability of finding an infected animal declines as the prevalence (x-axis) declines – starting at 100% and decreasing. The 95% probability level is highlighted by the horizontal line.

Updating the results in Fig 1 by bringing in results from previous studies we can say with 95% confidence that if human infective trypanosomes were circulating, then the prevalence among all livestock was less than 1 / 2848; among cattle, the corresponding prevalence was less than 1 / 2369 and among pigs less than 1 / 479 (Fig 1).

A total of 140 samples were positive by PCR (prevalence 4.2%), and of these 40 were also positive by HCT out of a total of 63 positive by HCT (prevalence 1.9%). Among cattle samples, 60 were positive by HCT; 40 were also positive by PCR; while 20 were negative by PCR. Of the 40 positive by PCR, 5 were positive for *T. vivax*, 14 for *T. congolense*; 4 for brucei and 34 for *T. theileri,* a common but benign species in cattle transmitted by Tabanids. Noteworthy is that by PCR, 41 were positive for *T. theileri* and so of these, the majority (83%) were also positive by HCT; this is in contrast to *T. brucei* (22.2% positive by HCT), *T. congolense* (21.2% positive) and T. vivax (9.3% positive).

### Results in areas with vector control activities

Comparing the results from the livestock samples to the distribution of vector control activities, 60.4% of samples were from areas undergoing vector control, comprising a greater proportion of porcine samples, with 64.6% of pig samples and 59.6% of bovine samples from areas undergoing vector control.

Evaluating the results of different PCR tests by trypanosome species and the vector control status, there was no significant difference in any of the tests (p > 0.05), once clustering at the village level was accounted for in a mixed model.

## Discussion

This study set out to investigate the possible existence of potentially human infective trypanosomes circulating within domestic animals in the *T. b. gambiense* endemic districts of Northwestern Uganda, as a basis to inform gHAT elimination activities in the region. Presence of an undetected animal reservoir would frustrate elimination efforts as it would provide a source of infection from which the existing tsetse flies would continue transmitting gHAT. Previous studies conducted in West Africa had indicated that several animals could act as reservoirs of *T. b. gambiense* in countries such as Cameroon, Guinea, and Ivory coast, among others [25].

The results of this study were surprising in that 20 of the samples in which trypanosomes had been detected by the HCT did not yield any signals from PCR, which is supposed to be highly sensitive, yet it detected signals in samples in which trypanosomes were not detected by the HCT. This could have been due to failure of primers to adequately anneal at the target templates, or to limiting parasite numbers even though they had been detected by parasitology; the parasitaemia was generally low, in many cases observing just a single trypanosome wriggling in the buffy coat area. The very few detectable trypanosomes could have been lost during sample processing, leading to negative PCR results. In a separate study, Ilboudo et al. (2023) [26]observed that some parasitology positive samples from experimentally infected pigs could turn out negative by PCR at some time points, mirroring our observations in field collected samples. The main issue in this case is the high detection threshold of the HCT, known to be 500 trypanosomes/ml of blood [27] cited in [28]. Thus, samples with lower parasitaemia can be indicated as negative and yet be detected by the PCR. Further studies are required to elucidate this likely common occurrence in the field.

According to the ITS-1 PCR, most infections were *T. congolense* followed by *T. vivax* and *T. brucei* (Trypanozoon), and none of the *T. brucei* infections had any signals for *T. b. gambiense* or *T. b. rhodesiense*. The district veterinarians and farmers were sensitized about the need to deliberately control trypanosomiasis and tsetse flies even as a measure against the important animal disease affecting livestock productivity, while securing health by clearing any possible animal reservoir of HAT. Presence of the benign *T. theileri* indicates that the Tabanids are present alongside tsetse flies, with the associated impact on livestock, adding mechanical transmission of *T. vivax* [29] to biological transmission by tsetse flies. The presence of multiple infecting species could be expected in the natural setting in which tsetse flies obtain their blood meals from multiple hosts that could be carrying different trypanosome species.

In the gHAT focus of north-western Uganda, case numbers have been in decline, driven by integrated medical and vector control activities [2,5,6,30]. As a result of this reduction in incidence, the WHO in 2022 validated Uganda as having eliminated gHAT as a public health problem. As well as demonstrating low incidence in the human population, it was also necessary to demonstrate a reduction in numbers of vectors and the absence of human infective trypanosomes in the sizeable livestock population in this area.

As we were not able to detect human infective trypanosomes despite a relatively large sample size we concluded that if the pathogens were circulating then this would not have been above a very low prevalence. Whilst we cannot rule out a risk to humans, this is very unlikely when one considers the very poor vectorial capacity of tsetse flies and relatively low risk of onward transmission to humans, as demonstrated in modelling studies [31]. However, it must be noted that samples that were *T. brucei s.l.* but SRA-negative were presumed to be *T. b brucei* as per previous studies [10,32,33]. Furthermore, while evidence from this suggests no, or very little circulation of human infective trypanosomes in Ugandan livestock there are frequent movements of people and livestock from neighbouring countries, in particular gHAT endemic South Sudan.

This is in agreement with findings from a focus in Côte d’Ivoire, where much higher rates of infections with Trypanosome species found in all animals 30% of pig samples positive by PCR), but no samples were positive for *T. b. gambiense* [34]. However, in the same study, 27.6% of pig samples were positive by immunological testing using the trypanolysis test [34]. Noteworthy is a recent study in pigs which indicated that the trypanolysis test may be reactive even for non-*T. b. gambiense* infections [35]. In this study, trypanolysis was not done as it would not have given unequivocal evidence for presence of *T. b. gambiense* in the animals.

Previous studies have demonstrated that the deployment of Tiny Targets across NW Uganda has had significant impacts on the abundance of tsetse [3,5,36] which has resulted in reductions in the incidence of gHAT [4]. Modelling of vector control combined with screening-and-treatment of cases suggests that transmission of gHAT was interrupted in 2021 [37]. In accord with these earlier findings, we did not find any evidence that *T. b. gambiense* was present in the livestock population. Our findings also contribute empirical evidence surrounding the debate that livestock do not act a reservoir hosts for *T. b. gambiense* [25], which could prevent or delay the achievement of elimination of transmission.

Paradoxically, our findings did not demonstrate that there was a significant difference of the prevalence of *Trypanosoma* species that cause AAT between parishes where Tiny Targets were deployed or not. The absence of any significant effect may be because Tiny Targets reduce the populations beyond the areas where they are deployed [36]. The extensive impact is due to the mobility of tsetse and this phenomenon may be enhanced further by the mobility of livestock.

Stronger evidence for quantifying the impact of Tiny Targets on AAT might be inferred by comparison of the prevalence of AAT in livestock populations before and after Tiny Targets were deployed. Studies undertaken in 2013-14, before the large-scale deployment of Tiny Targets, estimated the prevalence of *T. brucei sl*. in cattle to be 1.8% (38/2088) [12] compared to 0.65% (18/2775) in this study. Comparing these two studies is severely limited since the geographical locations of the samples let alone the herds are not matched. Unfortunately, there are no comparable data for the prevalence of *T. vivax* and *T. congolense* in the earlier study.

### Strengths and limitations

One of the challenges of such analyses is reasonably measuring zero in a very large population. It is only possible to survey a small proportion of the livestock in the area. Hence, the approach that we adopted here was to demonstrate that the pathogen was circulating below a certain prevalence. The baseline rate that we have adopted here was 1/1,000 in that we have shown that the pathogen, if circulating, was circulating at a prevalence below 1/1,000. When augmented with previous studies the corresponding prevalence dropped to 1 / 2,848. Even if the pathogen were circulating at this very low prevalence the low number of flies and low vectorial capacity would give very little scope for livestock to contribute to transmission to human hosts [38]. It must be acknowledged that this was a survey of livestock and that any potential circulation in wildlife species was not investigated. Furthermore, the sampling was voluntary – based on farmers presenting their animals for testing - which was a pragmatic necessity. As we are looking at exposure to tsetse and trypanosomiasis we do not identify any systematic bias that could be introduced by this.

A major limitation of this study was the lack of an internal control to conform the presence of intact DNA (integrity). However, we believe that the failure of PCR on some samples was a result of limiting trypanosomal DNA as the parasitaemia in these samples was extremely low. We could detect 1 or 2 trypanosomes, commonly in just 1 out of 4 capillaries prepared per blood sample. Thus, under such circumstances, it is possible that the extracted blood aliquot could have missed sufficient numbers of trypanosomes to provide the DNA required for specific amplification. In their study, Cunningham et al. [12] showed that only around 60% of samples from cattle, pigs and tsetse had sufficient DNA to be detectable by species specific primers, pointing to the inherent challenge of the usually limiting parasitaemia in natural infections.

Another challenge is that the sensitivity of PCR in detecting T. b. gambiense can be low given the characteristically low parasitaemia. A more sensitive PCR test has been recently published to minimize this shortcoming [39], but it was not available at the time this study was executed. Nevertheless, we increased chances of detecting *T. b. gambiense* by doing the nested PCR version of Maina et al. [21], which in our hands always proved more sensitive than the conventional single PCR versions.

## Conclusions

Whilst this is the third study to measure levels of human infective trypanosomes in this focus it is an innovative study in that 1) we covered each of the districts in the focus (that existed at the time of the study) and 2) it was conducted following the implementation of vector control in the area which should have reduced transmission. In conclusion, this study has not demonstrated the presence of *T. b. gambiense* circulating in cattle and pigs from the endemic districts of Northwestern Uganda.

However, as a requirement of post elimination surveillance prescribed by the NTD elimination roadmap, surveillance should be continued, at least at the sentinel sites within the region, to pre-empt any possible re-emergence of gHAT as a public health problem.

## Data Availability

All data produced in the present work are contained in the manuscript

## Acknowledgements

We wish to thank the District Veterinary and Health Offices in the entire region for their support, the farmers who presented their animals for the surveys as well as all the technical staff for their various inputs.

## Author contributions

**Conceptualization.** Enock Matovu, Charles Wamboga, Steve Torr, Sylvain Biéler, Paul Bessell, Joseph Ndung’u

**Data Curation.** Enock Matovu, Annah Kitibwa, Darwin Bella Okot, Alex Boobo

**Formal analysis.** Paul Bessell

**Funding acquisition.** Sylvain Biéler, Joseph Ndung’u

**Investigation.** Enock Matovu, Annah Kitibwa, Darwin Bella Okot, Alex Boobo

**Methodology.** Enock Matovu, Charles Wamboga, Steve Torr, Sylvain Biéler, Paul Bessell, Joseph Ndung’u

**Supervision.** Enock Matovu, Charles Wamboga

**Visualisation.** Paul Bessell

**Writing – original draft preparation.** Enock Matovu, Sylvain Biéler, Paul Bessell, Joseph Ndung’u

**Writing – review & editing.** Enock Matovu, Annah Kitibwa, Darwin Bella Okot, Alex Boobo, Charles Wamboga, Steve J Torr, Sylvain Biéler, Paul Bessell, Joseph Ndung’u

---

**S1 File. Supplementary information file.** File containing supplementary tables and figures.

