## Supplementary material for "A survey of livestock to identify the presence of potentially human infective trypanosomes in the *Trypanosoma brucei gambiense* endemic districts of north-west Uganda": S1 File. Supplementary information file

Supplementary table S1 Laboratory results by species and village

| Village | District | Species | n | HCT | PCR | T.v | T.c | T.t | T.b | Vector control |
|---|---|---|---|---|---|---|---|---|---|---|
| Akua | Arua | Bovine | 192 | 6 | 4 | 0 | 1 | 4 | 2 | Yes |
| Akua | Arua | Porcine | 6 | 0 | 0 | 0 | 0 | 0 | 0 | Yes |
| Cokokobidi | Yumbe | Bovine | 217 | 5 | 11 | 5 | 1 | 4 | 1 | No |
| Esia | Adjumani | Bovine | 116 | 5 | 0 | 0 | 0 | 0 | 0 | No |
| Esia | Adjumani | Porcine | 43 | 0 | 0 | 0 | 0 | 0 | 0 | No |
| Gwere East | Moyo | Bovine | 161 | 1 | 3 | 0 | 2 | 1 | 0 | Yes |
| Gwere East | Moyo | Porcine | 72 | 0 | 0 | 0 | 0 | 0 | 0 | Yes |
| Hahua | Arua | Bovine | 179 | 1 | 1 | 0 | 0 | 1 | 0 | No |
| Hahua | Arua | Porcine | 81 | 0 | 0 | 0 | 0 | 0 | 0 | No |
| Jabala | Yumbe | Bovine | 181 | 3 | 3 | 0 | 3 | 3 | 1 | Yes |
| Kocia | Moyo | Bovine | 204 | 3 | 2 | 1 | 1 | 1 | 1 | No |
| Kocia | Moyo | Porcine | 37 | 0 | 0 | 0 | 0 | 0 | 0 | No |
| Koloa | Koboko | Bovine | 281 | 6 | 35 | 25 | 9 | 8 | 1 | Yes |
| Lorikowo | Amuru | Bovine | 249 | 6 | 16 | 4 | 11 | 3 | 3 | Yes |
| Lorikowo | Amuru | Porcine | 73 | 2 | 0 | 0 | 0 | 0 | 0 | Yes |
| Mindrabe | Koboko | Bovine | 155 | 1 | 41 | 14 | 29 | 1 | 8 | Yes |
| Nyatika | Koboko | Bovine | 193 | 2 | 2 | 1 | 0 | 1 | 0 | Yes |
| Obomiri | Yumbe | Bovine | 124 | 5 | 3 | 0 | 2 | 3 | 0 | No |
| Okutura | Amuru | Bovine | 184 | 8 | 12 | 2 | 4 | 6 | 1 | No |
| Okutura | Amuru | Porcine | 3 | 0 | 0 | 0 | 0 | 0 | 0 | No |
| Omgbo | Maracha | Bovine | 94 | 1 | 1 | 0 | 0 | 1 | 0 | Yes |
| Omgbo | Maracha | Porcine | 109 | 1 | 0 | 0 | 0 | 0 | 0 | Yes |
| Oninivu | Maracha | Bovine | 148 | 3 | 2 | 2 | 0 | 1 | 0 | Yes |
| Oninivu | Maracha | Porcine | 82 | 0 | 0 | 0 | 0 | 0 | 0 | Yes |
| Otubanga North | Moyo (Obongi) | Bovine | 97 | 4 | 4 | 0 | 3 | 3 | 0 | No |
| Otubanga North | Moyo (Obongi) | Porcine | 25 | 0 | 0 | 0 | 0 | 0 | 0 | No |

HCT = Haematocrit centrifugation technique; PCR = Polymerase chain reaction; T. v = s; T. c = *Trypanosoma congolense*; T. t = *Trypanosoma thileri*; T. b = *Trypanosoma brucei*

Supplementary Table S2. Results of mixed infections in PCR

| Species | Total |
|---|---|
| *T. congolense* & *T. thileri* | 11 |
| *T. congolense* & *T. brucei* | 7 |
| *T. vivax* & *T. thileri* | 5 |
| *T. vivax* & *T. congolense* | 4 |
| *T. congolense* & *T. thileri* & *T. brucei* | 3 |
| *T. thileri* & *T. brucei* | 1 |
| *T. vivax* & *T. brucei* | 1 |
| *T. vivax* & *T. congolense* & *T. brucei* | 1 |
| *T. vivax* & *T. congolense* & *T. thileri* & *T. brucei* | 1 |

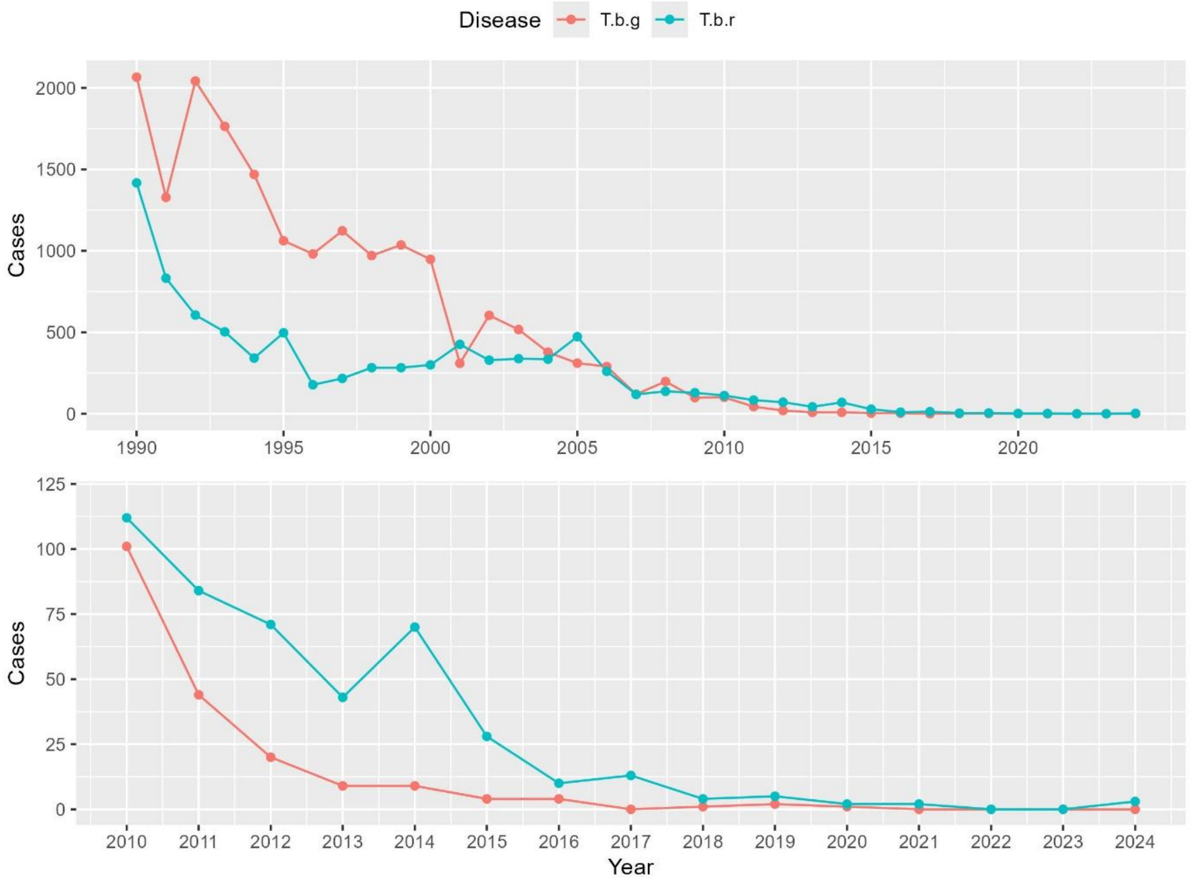


**Fig S1. Reported HAT cases in Uganda.** Top 1990-2023 and bottom 2010 – 2024 for the two species of human infective trypanosome – T.b.g = *Trypanosoma brucei gambiense*; T.b.r = *Trypanosoma brucei rhodesiense*. Data from WHO Global Health Observatory https://www.who.int/data/gho/data/themes/topics/human-african-trypanosomiasis.

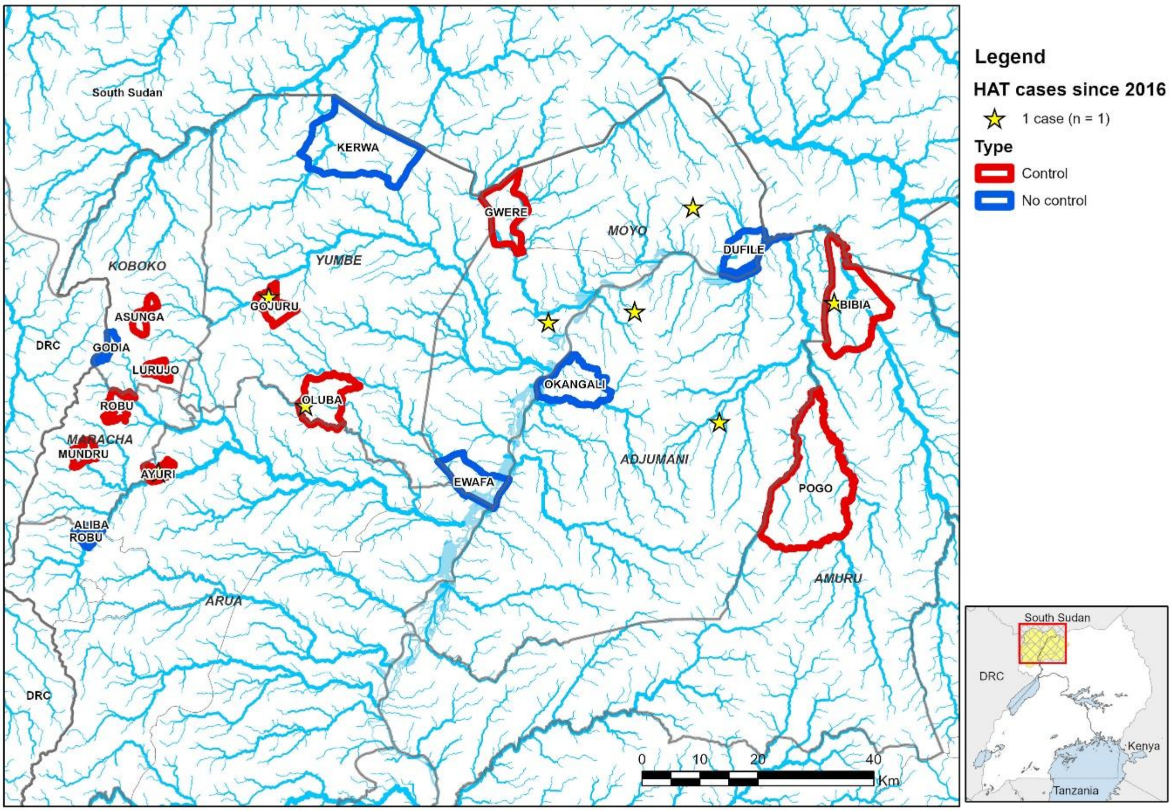


**Fig S2. Study area**. Study parishes included in the livestock survey based on whether there was ongoing tsetse control activities in the area. The stars denote cases that were detected between 2016 and 2018. Boundary data are from GADM (https://gadm.org/), rivers are derived by the authors from the NASA Shuttle Radar Topography Mission (SRTM) using the HydroSHEDS method [17]. Data are available from CC-BY compatible sources, The GADM licence is available at https://gadm.org/license.html, NASA STRM licence https://www.earthdata.nasa.gov/engage/open-data-services-software-policies/data-use-guidance.

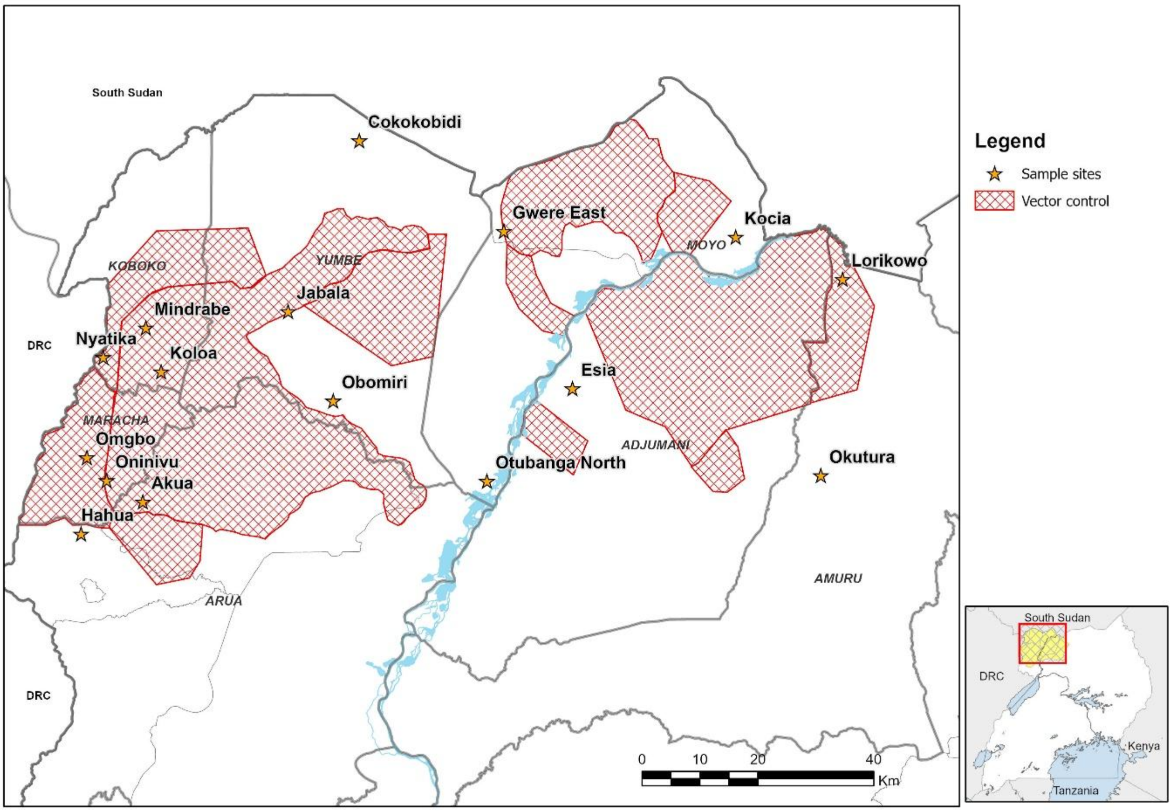


**Fig S3. Sampling sites.** Map of livestock sampling sites that were used in these analyses. The boundary and waterbody layers were obtained from CC-BY License compatible sources: GADM (http://www.gadm.org), and EnergyData (https://energydata.info/).Other layers were digitised by the authors. The GADM licence is available at https://gadm.org/license.html, Uganda waterbodies https://energydata.info/dataset/africa-water-bodies.
